# The contribution of malaria and sickle cell disease to anaemia among children aged 6 to 59 months in Nigeria: A secondary analysis using data from the 2018 Demographic and Health Survey

**DOI:** 10.1101/2022.03.23.22272825

**Authors:** Dennis L. Chao, Assaf P. Oron, Guillaume Chabot-Couture, Alayo Sopekan, Uche Nnebe-Agumadu, Imelda Bates, Frédéric B. Piel, Obiageli E. Nnodu

## Abstract

**Introduction:** Anaemia is a major cause of morbidity and mortality among children in sub-Saharan Africa. Anaemia has many aetiologies best addressed by different treatments, so regional studies of the aetiology of anaemia may be required.

**Methods:** We analysed data from Nigeria’s 2018 Demographic and Health Survey (DHS) to study predictors of anaemia among children ages 6-59m. We computed the fraction of anaemia at different degrees of severity attributable to malaria and sickle cell disease (SCD) using a regression model adjusting for demographic and socioeconomic risk factors. We also estimated the contribution of the risk factors to haemoglobin concentration.

**Results:** We found that 63.7% (95% CI: 58.3-69.4) of semi-severe anaemia (<80 g/L) was attributable to malaria compared to 12.4% (95% CI: 11.1-13.7) of mild-to-severe (adjusted haemoglobin concentration <110 g/L) and 29.6% (95% CI: 29.6-31.8) of moderate-to-severe (<100 g/L) anaemia and that SCD contributed 0.6% (95%CI: 0.4-0.9), 1.3% (95% CI: 1.0-1.7), and 7.3% (95%CI: 5.3-9.4) mild-to-severe, moderate-to-severe, and semi-severe anaemia, respectively. Sickle trait was protective against anaemia and was associated with higher haemoglobin concentration compared to children with normal haemoglobin (HbAA) among malaria-positive but not malaria-negative children.

**Conclusion:** This approach used offers a new tool to estimate the contribution of malaria to anaemia in many settings using widely available DHS data. The fraction of anaemia among young children in Nigeria attributable to malaria and SCD is higher at more severe levels of anaemia. Prevention of malaria and SCD and timely treatment of affected individuals would reduce cases of severe anaemia.

## Introduction

Anaemia is a major cause of morbidity and mortality among children. According to Global Burden of Disease (GBD) models [http://ghdx.healthdata.org/gbd-results-tool], there were 8.3 million (95% CI: 5.5-12.1M) years lived with disability (YLDs) worldwide among children under 5 years old due to anaemia in 2019, the majority of which (7.8M, 95% CI: 5.2-11.5M) were due to moderate or severe anaemia. The highest rates of anaemia (adjusted haemoglobin concentration <110 g/L) and severe anaemia (<70 g/L) among preschool-aged children are reported from sub-Saharan Africa (SSA) [1,2].

Anaemia can have many causes, such as nutritional deficiencies, infections, and haemoglobinopathies. In low-to-middle income countries (LMICs), children may have several concurrent causes for their anaemia, each requiring a different treatment strategy, for example iron supplementation, chemotherapy, transfusion, antimalarials, or deworming [3,4]. Iron deficiency is one of the largest causes of anaemia [2], but studies estimating the proportion of anaemia due to iron deficiency have highly variable results partly because diagnosing iron deficiency is difficult. A meta-analysis of 23 national surveys found that 25% of anaemia among pre-schoolers is associated with iron deficiency with high heterogeneity (range 0.3 - 51%) [5]. The gold standard test for diagnosis of iron deficiency, staining of a bone marrow sample for iron, is impractical as a screening method. Serum ferritin is commonly used as a marker of body iron stores, but it is an acute phase protein and therefore increases in the presence of inflammation, which is common in people living in LMICs [6].

Nigeria is Africa’s most populous country and experiences a high childhood anaemia burden. Previously, 60% of Nigeria’s moderate and severe anaemia among children under 5 years old has been attributed to iron deficiency and only 12% and 2% to malaria and sickle-cell disorders, respectively [http://ghdx.healthdata.org/gbd-results-tool]. Because most studies of anaemia are based on relatively small populations, national and regional estimates often need to extrapolate data from these geographically scattered studies to cover the rest of the population. Here, we have explored how Nigeria’s 2018 Demographic and Health Survey (DHS), a large, and uniquely detailed data source, could shed light on the contribution of *Plasmodium falciparum (P*.*f*.) malaria and SCD to anaemia. For the first time in DHS history, this Nigerian survey not only tested a population-representative sample of children for malaria infection, but also tested them for haemoglobin subunit beta (HBB) type, including the alleles associated with sickle cell trait and disease [7]. These biomarkers, plus the demographic information obtaineds from household interviews provide data on over 11,000 children ages 6 to 59 months. By modelling the risk of anaemia associated with these conditions, while adjusting for key demographic (age, sex) and socio-economic (household wealth, urban vs rural residence, maternal education) risk factors, we can provide a better estimate of the contribution of malaria and SCD to anaemia and explore the inter-relationships between SCD and trait, malaria, and anaemia.

## Data and Methods

### Data

We obtained data from Nigeria’s 2018 DHS [7]. A sample of 42,000 households were selected for participation in the survey. DHS interviews were conducted from August 14 to December 29, 2018. One-third of households (14,000) were randomly selected for biomarker evaluation. Children 6 to 59 months old who were *de facto* residents of these households (i.e., slept in household last night) were eligible for biomarker testing (11,536 eligible children). Blood samples taken from a finger or heel prick were analysed with a HemoCue analyzer in the field to determine haemoglobin concentration, which was adjusted for altitude, resulting in 11,206 (97.1% of eligible children) valid haemoglobin concentration measurements. The same finger or heel prick was used to evaluate malaria infection using SD Bioline AG *P*.*f*. (HRP-II) malaria RDT, yielding 11,173 (96.9% of eligible children) rapid diagnostic test (RDT) results. The same finger or heel prick was also used to evaluate HBB type using SickleSCAN point-of-care test from BioMedomics, yielding 11,186 results (97.0% of eligible children). This test determined the presence of the HbS and HbC alleles, allowing us to determine which children had normal HBB (HbAA), SCD (HbSS or HbSC), sickle cell trait (HbAS), or haemoglobin C trait (HbAC). The 9 children who had a recorded HBB type of “other” were excluded. In total, 11,142 children (96.6% of eligible children) with all three biomarker measurements were included in our analyses. The survey was approved by the National Health Research Ethics Committee of Nigeria and the ICF Institutional Review Board.

The DHS includes sample weights based on the survey design and on household response rates [7]. Applying the weights, the 11,142 children in our analysis represent 11,315.9 children. We applied sample weights for tabulating the number of children and for fitting regression models.

### Methods

We studied the relationships between anaemia and other factors using three thresholds for adjusted haemoglobin concentration: <110 g/L (anaemia), <100 g/L (moderate-to-severe anaemia), and <80 g/L (semi-severe anaemia) [8]. The threshold for severe anaemia among children is <70 g/L, but the less stringent threshold of 80 g/L instead of 70 was found to be a useful threshold for malaria-related anaemia [9]. We also found this cut-off allowed for identifying malaria RDT-negative children with haemoglobin C trait (HbAC) or SC disease (HbSC) in the DHS (Table 1).

**Table 1.** Anaemia severity by malaria RDT positivity and sickle cell result. The number of children ages 6 to 59 months, accounting for survey sample weights, is shown for each combination of RDT status, sickle cell genotype, and adjusted haemoglobin concentration. In each row, the percent of children in each anaemia severity category is shown in parentheses. An additional category of <80g/L, which covers all children with severe anaemia and the more severe end of moderate anaemia, is presented in the rightmost column.

|  |  | Moderate-to-severe anaemia (<100 g/L) |  | Mild or no anaemia (≥100 g/L) |  | **TOTAL** | **<80 g/L (semi-severe)** |
| --- | --- | --- | --- | --- | --- | --- | --- |
|  |  | <70g/L (severe) | 70-99g/L (moderate) | 100-109 g/L (mild) | ≥110 g/L (not anaemic) |  |  |
| RDT- | HbAA | 35.3 (0.6%) | 1441.8 (25.9%) | 1701.4 (30.6%) | 2386.0 (42.9%) | 5564.4 (100%) | 111.0 |
|  | HbAS | 6.1 (0.4%) | 467.9 (32.4%) | 420.4 (29.1%) | 549.4 (38.1%) | 1443.9 (100%) | 34.8 |
|  | HbAC | 0.0 (0.0%) | 38.1 (36.1%) | 30.2 (28.7%) | 37.2 (35.3%) | 105.6 (100%) | 1.3 |
|  | HbSS | 24.4 (32.3%) | 43.3 (57.4%) | 7.0 (9.3%) | 0.7 (1.0%) | 75.5 (100%) | 44.8 |
|  | HbSC | 0.0 (0.0%) | 13.8 (42.4%) | 16.3 (49.8%) | 2.6 (7.8%) | 32.6 (100%) | 2.6 |
| RDT+ | HbAA | 228.5 (7.2%) | 1794.3 (56.5%) | 655.9 (20.7%) | 497.5 (15.7%) | 3176.2 (100%) | 535.1 |
|  | HbAS | 30.4 (3.8%) | 442.1 (55.6%) | 183.3 (23.0%) | 139.9 (17.6%) | 795.7 (100%) | 90.7 |
|  | HbAC | 0.7 (0.9%) | 49.3 (61.9%) | 14.1 (17.7%) | 15.5 (19.5%) | 79.7 (100%) | 9.3 |
|  | HbSS | 12.5 (51.0%) | 10.9 (44.4%) | 1.1 (4.6%) | 0.0 (0.0%) | 24.5 (100%) | 18.3 |
|  | HbSC | 2.7 (15.3%) | 12.7 (71.0%) | 2.5 (13.7%) | 0.0 (0.0%) | 17.9 (100%) | 4.9 |
| **TOTAL** |  | 340.6 (3.0%) | 4314.3 (38.1%) | 3032.1 (26.8%) | 3628.8 (32.1%) | 11315.9 |  |

All the demographic and socio-economic data came from the DHS. To compare demographic characteristics between children with and without moderate-to-severe anaemia, we applied the Wilcoxon rank-sum test to determine if the distribution of ages was significantly different and a proportions test for the other characteristics (sex, malaria RDT positivity, HbAS, HbSS, rural residence, household in top wealth quintiles, and advanced maternal education).

We fitted multivariate quasibinomial logistic and multivariate linear generalized linear models (GLMs) to predict anaemia status and haemoglobin concentration, respectively. We used the svyglm package to fit these models using survey weights [https://r-survey.r-forge.r-project.org/survey/html/svyglm.html]. The primary sampling unit is the cluster (e.g., a village) and the strata are the DHS-defined strata (the states of Nigeria divided into urban and rural components).

We estimated the adjusted fraction of anaemia attributable to malaria by estimating the number of children with anaemia if no one had malaria using the fitted logistic regression model described above [10]. To construct the counterfactual simulation where no child is positive for malaria, we used the multivariate logistic regression models fitted to the original data to predict anaemia outcome for eligible children in the DHS after setting all of their malaria RDT results to negative. We interpreted the sum of the probabilities of these children being anaemic to be the number anaemic in the absence of malaria. Among children who were originally RDT positive, the sum of probabilities in the counterfactual population is interpreted to be the number of RDT-positive children who have anaemia that is not malaria-associated. We estimated the fraction attributable to SCD (HbSS/HbSC) by predicting the anaemia status of these children as if they had HbAA.

We computed the 95% confidence intervals of attributable fractions using the bootstrap. For each stratum in the DHS data, we drew clusters with replacement. Sampling by cluster instead of by individual preserved the clustering of children within households and households within villages [11,12]. We performed this procedure 1000 times to generate 1000 populations. We fitted models to these populations to compute the distributions of attributable fractions.

All analyses were performed in R version 4.0.5.

## Results

### Prevalence of anaemia

Among children aged 6 to 59m (N=11,142, or 11,315.9 with sample weights), 67.9% were anaemic (adjusted haemoglobin concentration <110 g/L), 41.1% had moderate or severe anaemia (<100 g/L), 7.5% had semi-severe anaemia (<80 g/L), and 3.0% had severe anaemia (<70 g/L) (Table 1). Children who were malaria RDT positive (RDT+) had higher prevalences of severe, semi-severe, and moderate-to-severe anaemia than those who were negative (Table 1). Of the children with semi-severe anaemia, 77.2% were RDT+, 7.4% had HbSS, and 82.4% were either RDT+ or had HbSS (Table 1). Among RDT+ children, 16.8% of those with HbAA and 11.4% of those with HbAS had semi-severe anaemia (Table 1). Severe anaemia was more prevalent among younger children except for those with HbSS (Figure S1). RDT positivity increased with age (Figure S2).

Children with moderate-to-severe anaemia were significantly younger and had lower proportions of females, malaria RDT+, HbSS, rural residence, lower 3 wealth quintiles, and lower maternal education compared to children with mild-to-no anaemia (Table 2).

**Table 2.** Characteristics of children with moderate-to-severe anaemia. The top row summarizes the mean age of children with moderate-to-severe or mild-to-no anaemia, and rows below show the average percentage of children within each anaemia category. Figures were computed without sample weights.

|  | Moderate-to-severe anaemia (<100g/L) | Mild or no anaemia (>=100g/L) | P value |
| --- | --- | --- | --- |
| N (unweighted) | 4660 | 6482 | --- |
| Mean age in years | 2.5 | 2.8 | <0.001 |
| % female | 46.4% | 51.6% | <0.001 |
| % RDT+ | 57.5% | 24.0% | <0.001 |
| % with HbAS | 20.2% | 19.2% | 0.19 |
| % with HbSS | 2.0% | 0.2% | <0.001 |
| % rural residence | 67.5% | 56.2% | <0.001 |
| % in top 2 wealth quintiles | 28.2% | 44.3% | <0.001 |
| % maternal education secondary school or greater | 33.3% | 46.9% | <0.001 |

### Predictors of anaemia

Malaria RDT positivity, HBB type, age, and wealth quintile were significantly associated with all degrees of anaemia (Table 3). Among children with HbAA, malaria positivity was associated with increased risk of mild-to-severe (4.05, 95% CI: 3.49-4.71), moderate-to-severe (5.06, 95% CI: 4.51-5.68), and semi-severe (9.11, 95% CI: 6.86-12.10) anaemia (Table 3). Among malaria-negative children, HbSS was associated with substantially increased anaemia risk compared to HbAA: an odds ratio of 69.66 (95% CI: 9.63-503.87) for mild-to-severe, 22.93 (95% CI: 10.56-49.78) for moderate-to-severe anaemia, and 77.36 (95% CI: 40.86-146.47) for semi-severe anaemia. We were unable to estimate the risk of mild-to-severe anaemia among RDT+ children with HbSS or HbSS because all these children had mild-to-severe anaemia (Table 1, Table 3). Although HbAS was associated with a slight increase in anaemia risk among RDT-children (1.19, 1.32, and 1.20 for mild-to-severe, moderate-to-severe, and semi-severe anaemia, respectively), HbAS was protective against anaemia among RDT+ children relative to HbAA (3.39/4.05=0.83, 4.23/5.06=0.84, and 5.28/9.11=0.58 for mild-to-severe, moderate-to-severe, and semi-severe anaemia, respectively).

**Table 3.** Predictors of anaemia. Multivariate logistic regression model for predicting mild-to-severe (left columns), moderate-to-severe (middle columns), and semi-severe anaemia (right columns). The effect of risk factors expressed as odds ratios.

|  | *Adjusted haemoglobin <110 g/L (anaemia)* |  |  | *Adjusted haemoglobin <100 g/L (moderate-to-severe anaemia)* |  |  | *Adjusted haemoglobin <80 g/L (semi-severe anaemia)* |  |  |
| --- | --- | --- | --- | --- | --- | --- | --- | --- | --- |
|  | *Odds ratio* | *(2.5%, 97.5%)* | *p* | *Odds ratio* | *(2.5%, 97.5%)* | *p* | *Odds ratio* | *(2.5%, 97.5%)* | *p* |
| *age in years* | 0.68 | (0.65, 0.71) | <0.001 | 0.71 | (0.69, 0.74) | <0.001 | 0.72 | (0.67, 0.77) | <0.001 |
| *female* | 0.86 | (0.77, 0.96) | <0.01 | 0.84 | (0.76, 0.93) | <0.001 | 0.89 | (0.74, 1.06) | 0.19 |
| *HBB/malaria status* |  |  |  |  |  |  |  |  |  |
| *HbAA/- (ref)* | -- |  |  | -- |  |  | -- |  |  |
| *HbAS/-* | 1.19 | (1.00, 1.41) | 0.04 | 1.32 | (1.13, 1.55) | <0.001 | 1.20 | (0.77, 1.88) | 0.42 |
| *HbAC/-* | 1.29 | (0.77, 2.17) | 0.34 | 1.43 | (0.86, 2.38) | 0.17 | 0.65 | (0.15, 2.78) | 0.56 |
| *HbSC/-* | 12.05 | (1.77, 81.82) | 0.01 | 2.43 | (0.58, 10.14) | 0.22 | 6.09 | (1.76, 21.09) | <0.01 |
| *HbSS/-* | 69.66 | (9.63, 503.87) | <0.001 | 22.93 | (10.56, 49.78) | <0.001 | 77.36 | (40.86, 146.47) | <0.001 |
| *HbAA/+* | 4.05 | (3.49, 4.71) | <0.001 | 5.06 | (4.51, 5.68) | <0.001 | 9.11 | (6.86, 12.10) | <0.001 |
| *HbAS/+* | 3.39 | (2.67, 4.30) | <0.001 | 4.23 | (3.49, 5.12) | <0.001 | 5.28 | (3.68, 7.57) | <0.001 |
| *HbAC/+* | 2.90 | (1.61, 5.23) | <0.001 | 4.95 | (3.03, 8.10) | <0.001 | 5.57 | (2.27, 13.67) | <0.001 |
| *HbSC/+* | * | * | <0.001 | 22.93 | (5.10, 103.13) | <0.001 | 38.76 | (10.78, 139.39) | <0.001 |
| *HbSS/+* | * | * | <0.001 | 62.43 | (13.83, 281.76) | <0.001 | 171.84 | (57.35, 514.89) | <0.001 |
| *rural (urban ref)* | 0.99 | (0.85, 1.14) | 0.86 | 0.92 | (0.80, 1.05) | 0.22 | 1.11 | (0.87, 1.41) | 0.40 |
| *wealth* |  |  |  |  |  |  |  |  |  |
| *richer/richest (ref)* | -- |  |  | -- |  |  | -- |  |  |
| *middle* | 1.03 | (0.88, 1.20) | 0.71 | 1.09 | (0.93, 1.28) | 0.28 | 1.06 | (0.80, 1.42) | 0.68 |
| *poorer* | 1.33 | (1.09, 1.63) | <0.01 | 1.34 | (1.11, 1.62) | <0.01 | 1.52 | (1.12, 2.07) | <0.01 |
| *poorest* | 1.72 | (1.36, 2.14) | <0.001 | 1.51 | (1.24, 1.84) | <0.001 | 1.81 | (1.31, 2.52) | <0.001 |
| *mother's education* |  |  |  |  |  |  |  |  |  |
| *none or primary (ref)* | -- |  |  | -- |  |  | -- |  |  |
| *Secondary or higher* | 0.85 | (0.73, 1.00) | <0.05 | 0.85 | (0.74, 0.98) | 0.03 | 0.81 | (0.64, 1.01) | 0.07 |

\* indicates odds ratios >1000.

### Anaemia attributable to malaria and SCD

We found the fraction of anaemia attributable to malaria to increase with the severity of anaemia, from 12.4% (95% CI: 11.1-13.7) of overall anaemia (adjusted haemoglobin concentration <110 g/L) to 29.6% (95% CI: 29.6-31.8) of moderate-to-severe anaemia (<100 g/L) and to 63.7% (95% CI: 58.3-69.4) of semi-severe anaemia (<80 g/L) (Figure 1A). The fraction attributable to SCD was 0.6% (95%CI: 0.4-0.9) of overall anaemia, 1.3% (95% CI: 1.0-1.7) of moderate-to-severe, and 7.3% (95%CI: 5.3-9.4) of semi-severe anaemia (Figure 1B).

**Figure 1.**
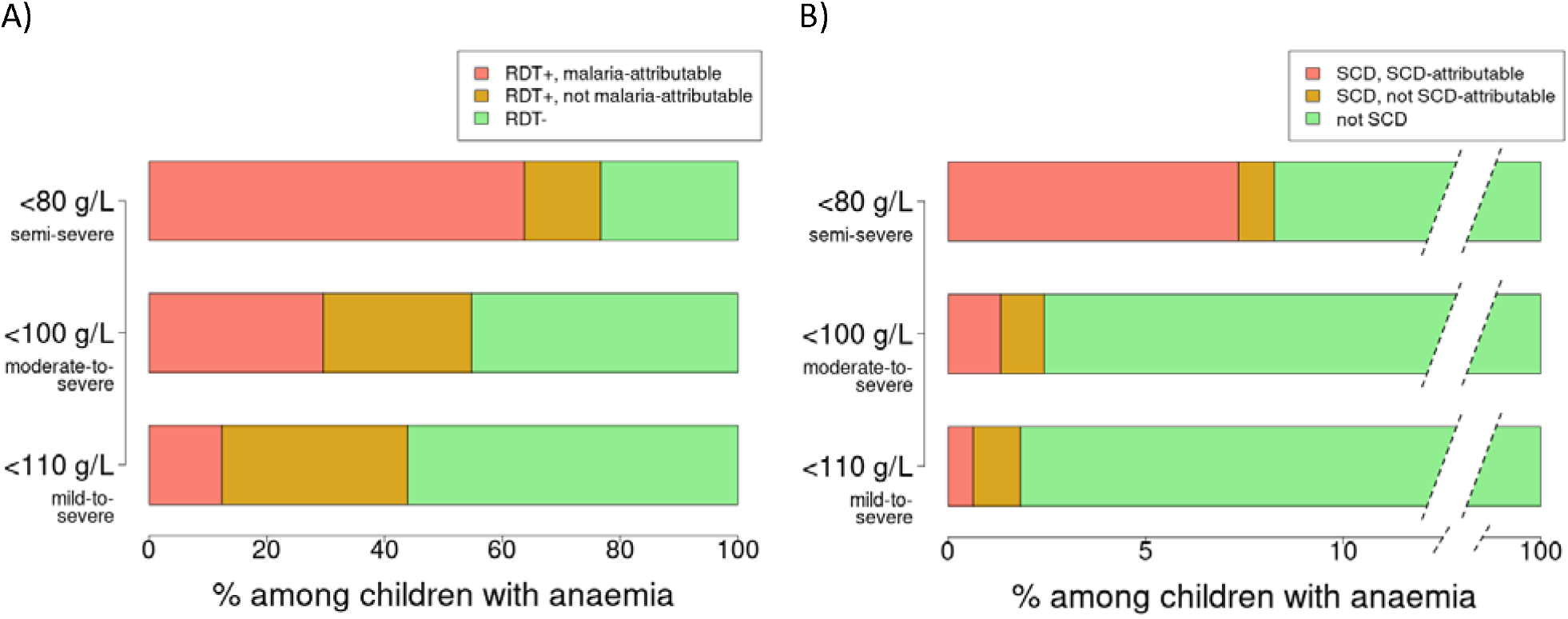
Fraction of anaemia attributable to malaria or SCD. Anaemia is defined as having an adjusted haemoglobin concentration of <110 g/L (bottom bars), <100 g/L (middle bars), or <80 g/L (top bars). A) The bars represent the fraction of these three levels of anaemia that can be malaria-attributed (anaemic children who were RDT+ but would not have been anaemic if they had been RDT-according to our model), RDT+ but not malaria-attributed (RDT+ children who would have been anaemic even if they were RDT-), and RDT-. B) The fraction of anaemic children that can be attributed to SCD.

Although RDT positivity is a major risk factor for anaemia at all levels of severity (Table 3), malaria might not be the cause of anaemia among all children who were RDT+. Among RDT+ children with anaemia, we estimated that 69.3% (95% CI: 71.8-74.2) would still have been anaemic if they had been RDT-, 46.0% with moderate-to-severe anaemia (95% CI: 43.3-48.7) would have had moderate-to-severe anaemia had they been RDT-, and 17.8% with semi-severe anaemia (95% CI: 13.5-20.6) would have had semi-severe anaemia even if they were RDT-.

Children with HbSS or HbSC were at high risk of anaemia (Table 3), but some of these children would still have anaemia even if they did not have SCD. If these children with SCD had HbAA instead, 60.0% (95% CI: 65.8-70.7) of those who had anaemia would still have anaemia, and 45.2% (95% CI: 39.3-52.9) and 11.2% (95%CI: 8.1-14.6) with moderate-to-severe and semi-severe anaemia would still have those levels of anaemia.

Sickle trait (HbAS) was associated with protection against anaemia among malaria-positive children (Table 3). Our model predicts that if children with HbAS had HbAA instead, their prevalence of mild-to-severe and moderate-to-severe anaemia would be higher (69.1% vs 67.4% and 42.2% vs 40.0%), but semi-severe anaemia would increase from 5.4% to 7.3%, which is close to the 7.4% prevalence of semi-severe anaemia observed among children with HbAA when including RDT- and RDT+ children (Table 1).

If the information about HBB type is not used by the regression models used for prediction (Table S1), then the estimated fraction of anaemia attributable to malaria drops from 12.4 to 12.3%, moderate-to-severe anaemia attributable to malaria drops from 29.6 to 29.4%, and semi-severe from 63.7 to 62.2%.

### Predictors of haemoglobin concentration

Each year of age was associated with an increase in haemoglobin of 2.74 g/L (Table 4). Among children with HbAA, RDT positivity was associated with a 12.93 g/L lower haemoglobin concentration (Table 4). Among malaria negative children, haemoglobin concentration was 28.92 g/L lower among children with HbSS than HbAA. However, among RDT+ children, those with HbAS had higher haemoglobin concentration than HbAA (−10.76 - -12.93=2.17 g/L). Each of the lowest two wealth quintiles had lower haemoglobin concentrations than the combined top two and having a mother with a secondary school or higher education had higher haemoglobin concentrations than a mother with primary or no education (1.12, 95% CI: 0.28-1.97).

**Table 4.** Predictors of haemoglobin concentration. Multivariate linear regression results to predict adjusted haemoglobin concentration. The baseline haemoglobin concentration is in the first row, with the following rows summarizing the effect of risk factors.

|  | *g/L* | *(2.5%,97.5%)* | *p* |
| --- | --- | --- | --- |
| **Baseline concentration** | 99.29 | (98.06,100.52) | <0.001 |
| **age in years** | 2.74 | (2.50,2.97) | <0.001 |
| **Female (ref: male)** | 1.23 | (0.61,1.85) | <0.001 |
| **HBB/malaria status** |  |  |  |
| **HbAA/- (ref)** | -- |  |  |
| **HbAS/-** | -1.87 | (-2.80, -0.94) | <0.001 |
| **HbAC/-** | -1.15 | (-3.89, 1.58) | 0.41 |
| **HbSC/-** | -12.35 | (-15.90, -8.80) | <0.001 |
| **HbSS/-** | -28.92 | (-33.31, -24.52) | <0.001 |
| **HbAA/+** | -12.93 | (-13.76, -12.10) | <0.001 |
| **HbAS/+** | -10.76 | (-12.02, -9.49) | <0.001 |
| **HbAC/+** | -9.42 | (-12.65, -6.20) | <0.001 |
| **HbSC/+** | -27.85 | (-35.65, -20.06) | <0.001 |
| **HbSS/+** | -37.25 | (-43.56, -30.94) | <0.001 |
| **rural (ref: urban)** | -0.03 | (-0.87, 0.81) | 0.94 |
| **wealth** |  |  |  |
| **richer/richest (ref)** | -- |  |  |
| **middle** | -0.15 | (-1.05, 0.76) | 0.75 |
| **poorer** | -1.89 | (-3.04, -0.74) | <0.01 |
| **poorest** | -3.80 | (-5.10, -2.50) | <0.001 |
| **mother's education** |  |  |  |
| **none or primary (ref)** | -- |  |  |
| **Secondary or higher** | 1.12 | (0.28,1.97) | <0.01 |

## Discussion

We found that malaria RDT positivity and SCD were associated with a high risk of anaemia among children participating in Nigeria’s 2018 DHS. Our results support the hypothesis that malaria and SCD play a larger role in more severe compared to mild anaemia. Our estimate of the contribution of malaria to moderate-to-severe anaemia among young children in Nigeria of 30% is substantially higher than the GBD’s estimate of 12% [http://ghdx.healthdata.org/gbd-results-tool]. We found that higher levels of household wealth and maternal education were associated with reduced risk of anaemia and higher average haemoglobin concentration in children, which is consistent with earlier analyses of the same dataset [7,13] and more broadly in sub-Saharan Africa [14]. However, the effects of malaria RDT positivity and SCD on anaemia were much larger than these sociodemographic factors.

Testing for malaria infection could help assess the aetiology of anaemia in individuals, but when malaria prevalence is high, many children with anaemia from non-malarial causes could incidentally have asymptomatic or mild malaria. In our analysis, 77% of children with adjusted haemoglobin concentration <80 g/L were malaria-positive, but our model indicates that 18% of these children would have had anaemia even if they were not malaria-positive. At the less stringent <100 g/L threshold, 46% of malaria-positive children with moderate-to-severe anaemia would have had anaemia if they had not been malaria-positive, indicating that malaria is a larger contributor to more severe anaemia. These findings are important because they mean that anaemia in children with malaria cannot be assumed to be due to malaria and therefore the anaemia needs to be investigated and managed appropriately. Previous studies have found that a high proportion of children with apparent symptomatic malaria might have had a different underlying aetiology for their symptoms. An estimated 1/3 of a cohort of children hospitalized for severe malaria in Kenya may have had non-malarial illness [15]. Dalrymple et al 2019 estimated that only 37% of fevers among malaria-positive children in sub-Saharan Africa were due to malaria [16].

Although only 1.3% of children had SCD (HbSS or HbSC), our analysis attributes 7.3% of semi-severe anaemia (haemoglobin concentration <80 g/L) to SCD. We note that 10% of children with severe anaemia (<70 g/L) had HbSS or HbSC. Ideally all children with SCD should be identified through a newborn screening program. However, such programs are scarce in Africa but as an interim measure our findings indicate that, because SCD is so prevalent in our population-representative sample of children with severe anaemia, screening all young children with severe anaemia with a point-of-care test could identify undiagnosed SCD. Active management could then be started, which would improve their intellectual and physical development and could reduce their need for transfusions. Olupot-Olupot et al 2022 describe a risk-assessment algorithm that identified 73% of undiagnosed sickle cell anaemia among children admitted to the hospital for severe anaemia [17].

The prevalence of severe anaemia is substantially lower among the 20% of children with sickle trait (HbAS) than those with HbAA (1.6% vs 3.0%), which we assume to be due to protection against malaria. Although children with sickle trait had a slightly lower haemoglobin concentration than children with HbAA, sickle trait was associated with protection against anaemia among malaria-positive children, with a greater degree of protection when using the more stringent anaemia cutoff. This would be consistent with observations that sickle trait protects against severe malaria more than mild malaria illness [Williams et al 2005] and reduces hospital admissions for malaria (OR=0.26) [18]. Sickle trait did not appear to be associated with protection against getting malaria since children with HbAS did not have lower malaria RDT positivity [Figure S2 and 19]. Our estimate of the protection of HbAS against malaria-associated anaemia is generally consistent with the literature. Sickle cell trait may confer over 90% protection against severe malarial anaemia [4,20]. Lopera-Mesa et al 2015 found a clinical malaria incidence rate ratio of 0.66 between children with HbAS vs HbAA [21]. This effect was most apparent in early childhood, peaking around 3-6 years of age.

Our study has several limitations. Although we found that increasing age was associated with a reduced risk of anaemia, this finding could be partially attributed to the fixed thresholds used to define anaemia for all children regardless of age. Haemoglobin concentrations increased with age, which has been observed in healthy, disease-free children [22]. Using a single threshold for all ages could result in an over-estimate of anaemia among young children and an under-estimate at older ages. Of note, current anaemia thresholds were set in 1968 using mostly European and North American populations, and the WHO has begun a process to revise them [23]. The DHS interviews were conducted between August and December, which overlap with malaria season in northern Nigeria, and the estimates of malaria prevalence and its impact on anaemia could be different across seasons. We estimated the fraction of anaemia attributable to malaria and sickle cell disease using a simple statistical model, which would have been more accurate if assays for other major causes of anaemia, such as helminths, genetic conditions besides SCD, and iron or other micronutrient deficiency, were included. A combination of these tests would be key to determining the aetiology of an individual’s anaemia. We equated malaria RDT status with malaria, but the antigens detected by the RDT can persist for weeks after malaria treatment [24]. Similarly, only data on *P. falciparum* malaria were available in the DHS, not on P. vivax, though falciparum comprises most infections is in Nigeria. Even with additional data, the causes of anaemia are complex and can be interrelated, making it difficult to assign aetiology. Calis et al 2008 used structural equation modelling to account for these relationships [25]. We were limited to data available in the DHS, and our simpler approach was able to capture the relationship between malaria positivity, sickle trait, and sickle cell disease. We found that using the malaria test data alone (I.e., ignoring the HBB type) would slightly underestimate the fraction of anaemia attributable to malaria. Keeping this effect in mind, we believe that our approach can be applied to DHS datasets that do not include HBB typing to estimate the fraction of anaemia attributable to malaria.

Our analyses highlight the importance of malaria and SCD in the more severe stages of anaemia among young children in Nigeria. The majority of children in the DHS survey had mild-to-severe anaemia, most of which was not due to malaria or sickle cell disease. At the more severe end of the anaemia spectrum, malaria appears to be linked to 63.7% of semi-severe cases and sickle cell disease to 7.3%. There are many ways to prevent or mitigate anaemia early, which could avert the development of severe anaemia and the need for blood transfusions. Malaria control has been observed to reduce anaemia prevalence [26,27]. Mass iron supplementation or fortification is suggested for regions with high anaemia prevalence, but because iron supplementation can increase the susceptibility of malaria, malaria diagnosis, prevention, and treatment are important components of anaemia-reduction in malaria-endemic regions [28]. However, existing coverage of such anti-malaria measures is often inadequate for the safe provision of iron to children [27]. Abhulimhen-Iyoha et al 2018 found that the majority of blood transfusions required by children in a tertiary hospital in Nigeria were due to severe malaria and suggest that malaria prevention could alleviate pressure on the nation’s limited blood supply [29]. The early diagnosis and prophylactic treatment for SCD in certain populations could also be part of an anaemia-control strategy, a possibility made more feasible with the recent development of point-of-care tests for the condition [30]. We believe that a more complete and region-specific understanding of the aetiology of anaemia, particularly severe anaemia, is required to prioritize prevention efforts and assist diagnosis and appropriate treatment.

## Data Availability

The data used in this study can be obtained from the Demographic and Health Survey programme by registered users upon approval of a study description. Instructions for obtaining these data are at https://dhsprogram.com/.

## Contributors

DLC, APO, GCC, and OEN conceived the study. DLC and APO designed the study. DLC and APO performed the analyses and verified underlying data. DLC, APO, GCC, AS, UN-A, IB, FBP, and OEN interpreted the data and results. All authors participated in the writing of the manuscript.

## Acknowledgments

DLC and GCC are employees of the Bill & Melinda Gates Foundation. OEN is the primary investigator of the Sickle Pan African Research Consortium in Nigeria and acknowledges support from the National Heart, Lung, and Blood Institute for the Sickle Pan African Research Consortium (NIH U24HL135881). FBP acknowledges infrastructure support for the Department of Epidemiology and Biostatistics provided by the NIHR Imperial Biomedical Research Centre. We thank the Demographic and Health Survey programme and the National Population Commission and the Department of Health, Non-Communicable Diseases Unit, Federal Ministry of Health, Nigeria for making our work possible. We also thank Michelle O’Brien and Edward Wenger for helpful conversations.

## Supplemental table

**Table S1.**
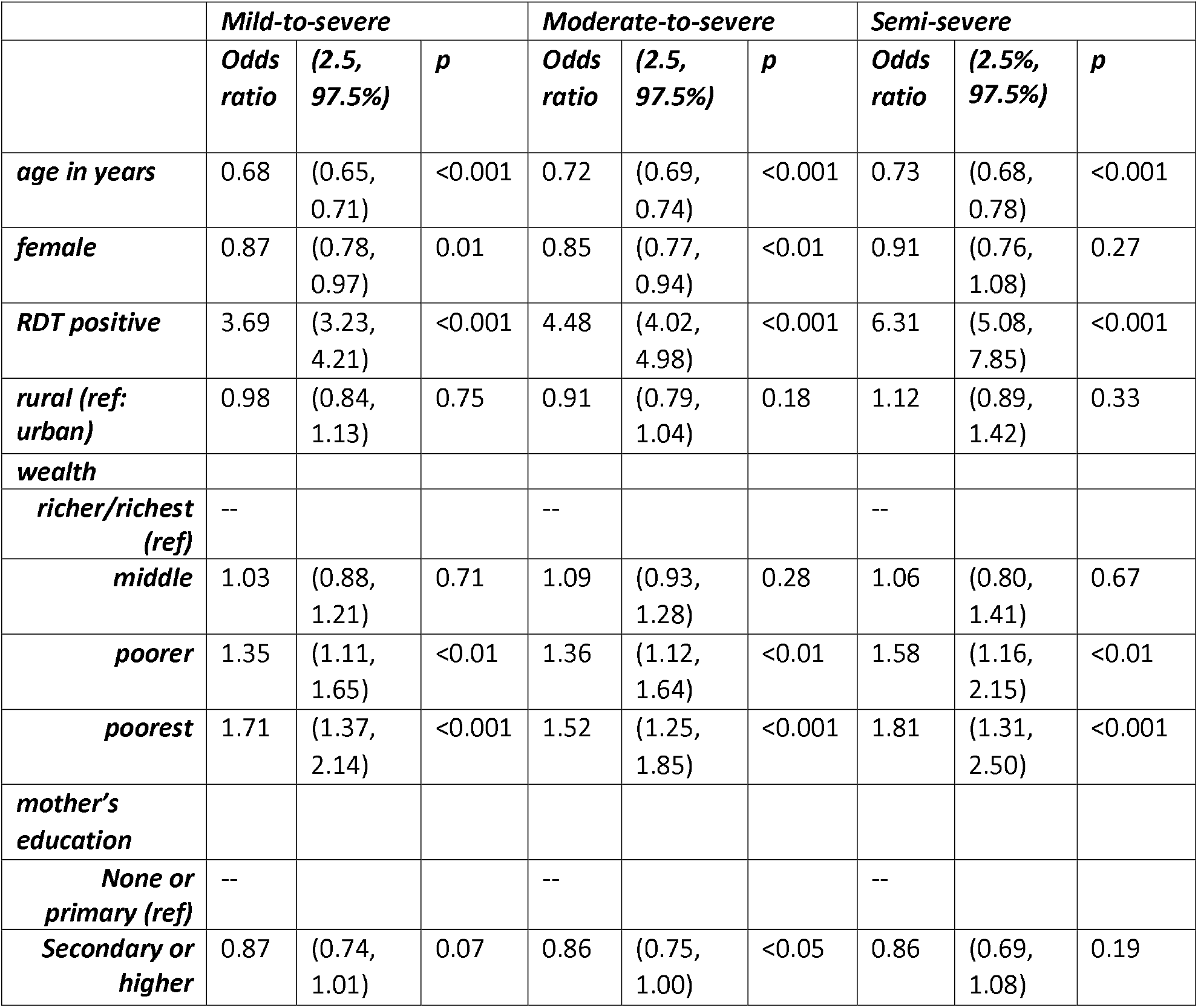
Predictors of mild-to-severe, moderate-to-severe, and semi-severe anemia without using sickle cell status.

## Supplemental figures

**Figure S1.**
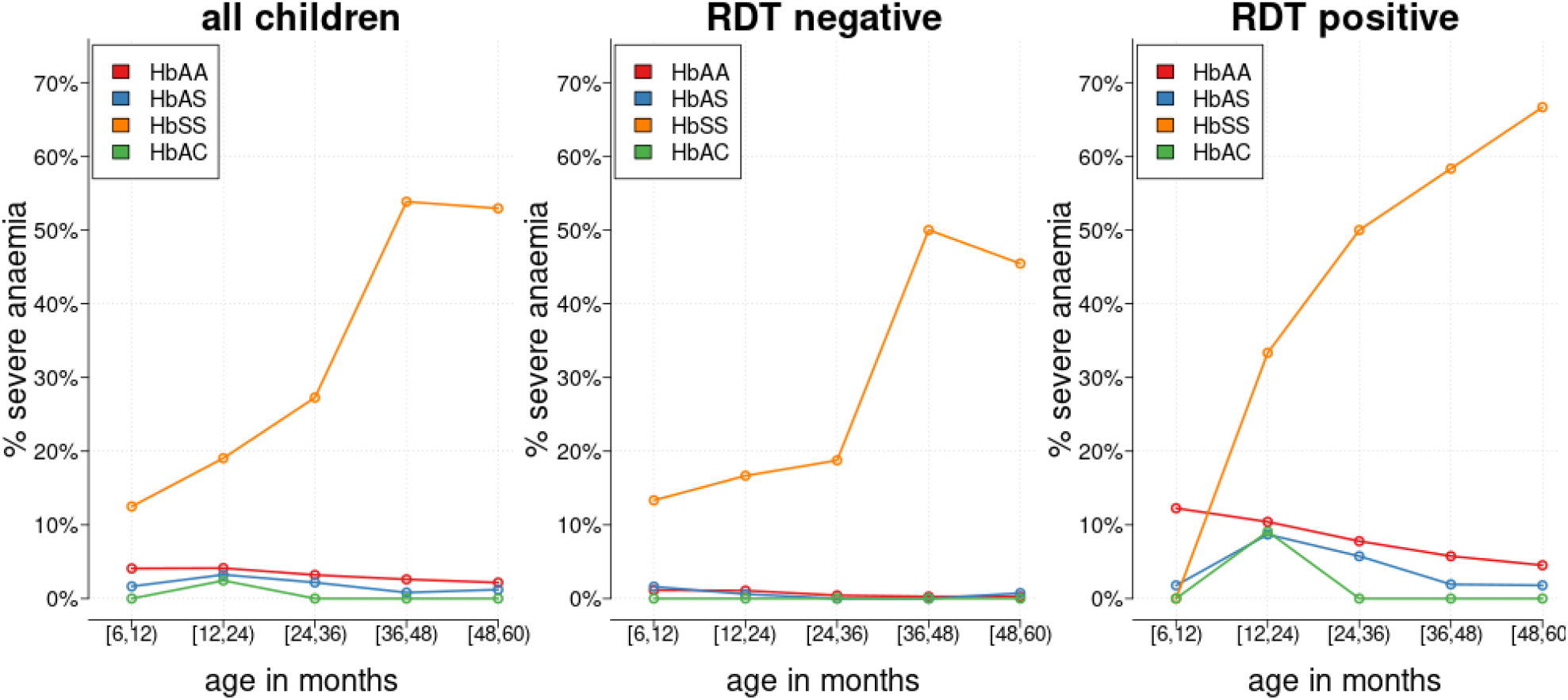
Severe anemia by age and sickle cell genotype. Among children with HbSS, the prevalence of severe anemia increases dramatically with age. Among children without HbSS, the prevalence of severe anemia decreases with age. Also note the difference between HbAA (red) and HbAS (blue) among RDT-positive children.

**Figure S2.**
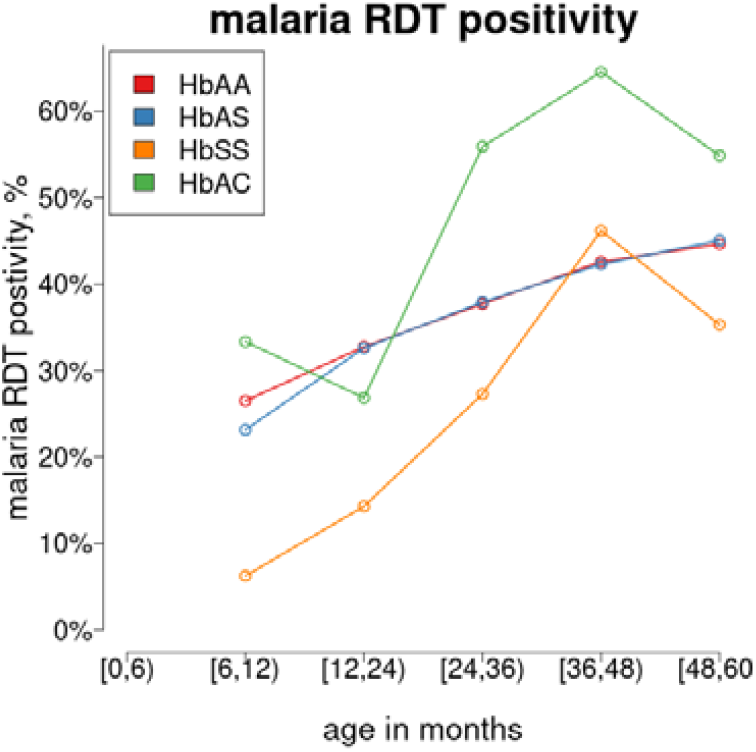
Malaria positivity by age and sickle cell genotype.

